# DEEP LEARNING-BASED PHENOTYPING OF FOREFOOT MORPHOLOGY IN HEREDITARY THORACIC AORTIC DISEASES

**DOI:** 10.1101/2025.02.06.25321829

**Authors:** Diego Ulisse Pizzagalli, Susanna Grego, Stefanos Demertzis

## Abstract

Hereditary thoracic aortic diseases (HTAD) are often associated with multifaceted phenotypic manifestations in different anatomical districts, including skeletal abnormalities. Therefore, diagnostic criteria account for multiple parameters to compute a systemic risk score. Despite the forefoot is known to be different in HTAD, its complex morphology is difficult to be quantified objectively and it is not currently considered in diagnostic criteria.

Here, we investigated the potential application of artificial intelligence to compute a HTAD risk score from smartphone-acquired images of the forefoot. To this end, we conducted a pilot study including 44 adults, of which 22 had high risk of HTAD (in line with EACTS/STS guidelines 2024 and revised Ghent criteria). The remaining 22 individuals did not show characteristic features indicative of HTAD. A deep learning architecture was then trained to compute a risk score using specific strategies to account for limited sample sizes: transfer learning and leave-one-out cross validation.

The computed risk score was significantly higher in the HTAD group with respect to the control group (p < 0.0001), achieving remarkable sensitivity (82%) and specificity (91%), with an AuC of 0.94.

Altogether this study highlights the usefulness of AI to assist the analysis of complex morphological traits, potentially enabling a greater number of healthcare professionals to identify patients at risk of HTAD and readily address them to a proper clinical examination.

The study was approved by the Swiss Cantonal Ethics Committee with protocol number 2023-00643.

## INTRODUCTION

Hereditary thoracic aortic diseases (HTAD) pose acute life-threatening complications such as aortic dissection and aneurism rupture ^1,2^. Their origin often involves variations in genes encoding for crucial components of the connective tissue and extracellular matrix, such as FBN1 in Marfan syndrome, one of the major causes of HTAD ^3–5^. Consequently, phenotypic manifestations are multifaceted and can affect multiple anatomical districts.

Clinical diagnosis therefore necessitates comprehensive criteria such as the revised Ghent nosology that allowed to compute a systemic risk score by the sum of multiple parameters including ocular, cardiovascular, and skeletal abnormalities ^6,7^. Amongst these, foot abnormalities such as hindfoot deformity and pes planus contribute with 2 and 1 points respectively but lack diagnostic sufficiency.

Despite the forefoot is known to be different in patients with HTAD, currently it is not included in criteria for disease risk evaluation. Indeed, its complex morphology, the large difference across individuals, and potential interobserver bias pose challenges for its quantification in the clinical practice.

Recently, artificial intelligence (AI) demonstrated promising applications for assisted patient evaluation, with the potential to be smoothly integrated into existing clinical workflows ^8^.

Therefore, we explored the usage AI for the automated analysis of the complex forefoot morphology. This enabled us to compute a risk score for HTAD directly from images of the forefoot captured via a smartphone with high sensitivity and specificity, supporting the association between forefoot abnormalities and HTAD, and providing a convenient methodology that could complement the existing clinical evaluation criteria.

## RESULTS

To evaluate the possibility of applying AI for convenient analysis of forefoot morphology we acquired smartphone-pictures along with cardiovascular examination from a pilot cohort of 44 individuals. (Fig. 1A). Then, the acquired images were cropped to include the metatarsophalangeal region, as this area exhibited less pose-dependent variability across subjects (Fig. 1B). Amongst the collected images, 22 were from patients with high risk of having hereditary thoracic aortic disease in line with EACTS/STS guidelines 2024 ^9^ displaying aortic root enlargement (z-score >= 2) and/or a systemic score > 7 according with Ghent revised criteria. The remaining 22 individuals did not show characteristic features indicative of HTAD, pathological aortic root dilation or aortic dissection (Fig.1 C).

**Figure 1.**
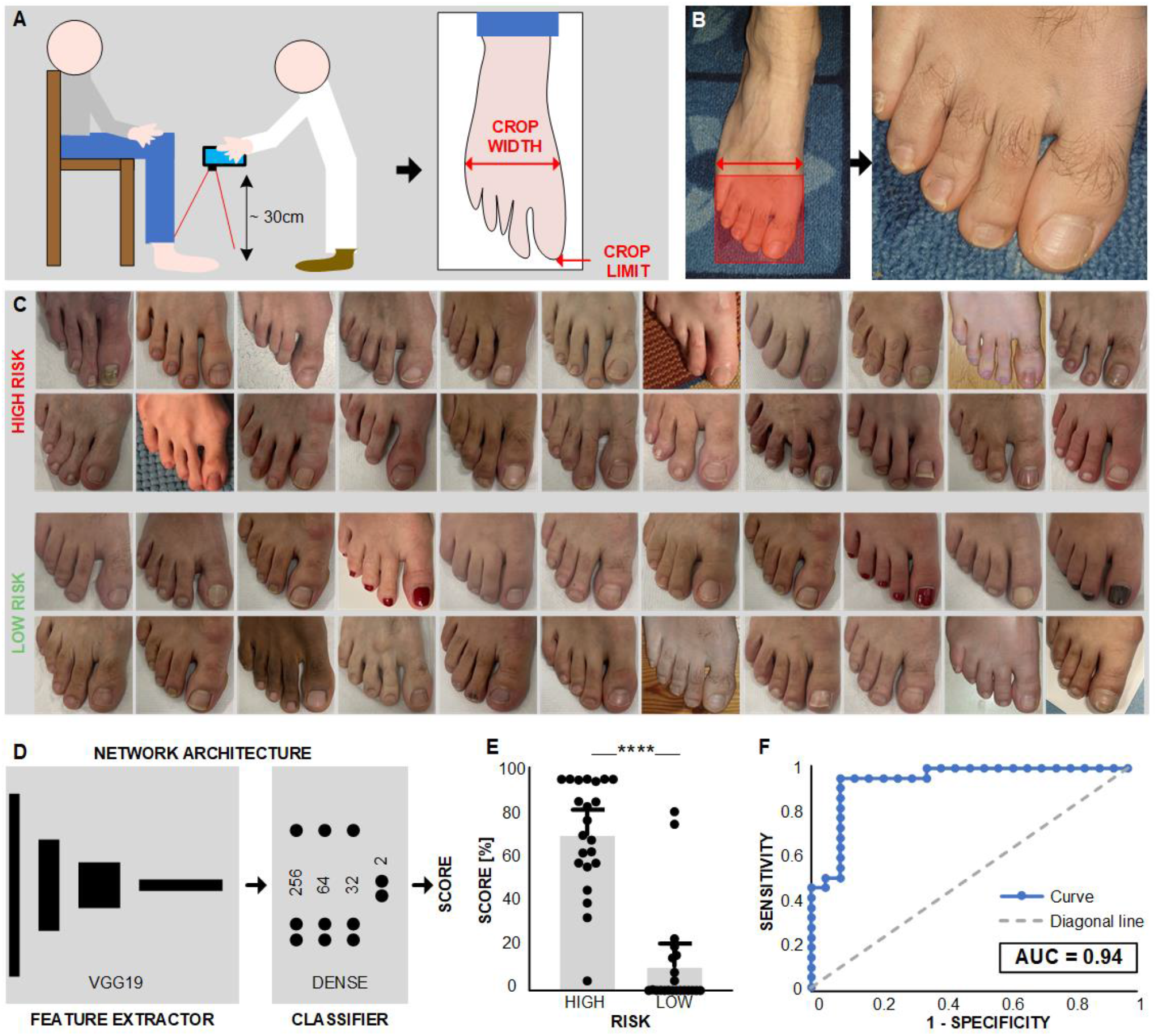
AI-driven phenotyping of foot morphology. **A**. Schematic representation of the data-acquisition process showing the capturing of pictures with the frontal view of the foot. **B**. Crop of the images to encompass the metatarsophalangeal region. **C**. Dataset used for training with images divided into two groups: high risk and low risk for HTAD. **D**. Deep learning architecture composed of a feature extractor block (VGG19), pretrained on ImageNet, and a classifier block composed of a three fully connected layers. **E**. Risk score computed via leave-one-out validation on foots from patients from two groups showing a significant difference (p < 0.0001) (n = 44 individuals. Bar plots indicate mean and 95% CI, p-values computed using unpaired t-test). **F**. Receiver Operator Curve (ROC) showing an area under the curve of 0.94.

Then, we trained a deep learning architecture composed of two blocks: a VGG19^10^ block used as feature extractor, and a custom morphology classifier block based on a fully connected network (Fig. 1D). Due to limited dataset size, we used a transfer learning approach, pre-training the VGG19 on the ImageNet dataset ^11^. Then, the architecture was trained on our dataset and validated using the leave-one-out method.

Results indicate remarkable sensitivity (82%) and specificity (91%), p-value < 0.0001 across the two groups, and AuC of 0.94 (Fig. 1E-F). Amongst four individuals in the high-risk group that received a low score (false negatives), three were affected by alternative mutations (MYLK1, and LDS syndrome). Conversely, one patient that received a high score but included in the low-risk group due to low systemic score and normal aortic root diameter, had a family history for aortic dissection.

## METHODS

Patient Recruitment and Data Acquisition: Patient recruitment followed approval by the Swiss Ethics Committee (Swiss ethics). All participants were provided with detailed information regarding purpose, rationale, and significance of the project, as well as data management procedures, and signed an informed consent. Participants were divided into high risk and low risk groups upon clinical examination that included computation of risk score in line with revised Ghent criteria and HTAD guidelines 2024, transthoracic ultrasound, familiarity and genetic testing when these data were available.

Images were captured with the patient in a seated position, with their foot resting on the floor. Only the right foot of each participant was included in the analysis. The images were taken from a superior-frontal perspective by a cardiologist using an iPhone SE 2020. The use of flash was disabled during photography to minimize potential glare or shadow artifacts. The distance between the camera and the foot was approximately 50 centimeters, ensuring consistent framing and focus across all images.

Deep Learning Pipeline for Forefoot Morphology Classification: Prior to processing, images were standardized by cropping (square crop, size: max. width of the foot, corner: right toe extremity), and subsequently rescaled to 224×224 pixels. Augmentation techniques were applied to diversify the training dataset, including rotation within ±10 degrees, zoom within ±15%, horizontal and vertical shifts within ±10%. Transfer learning was employed using the VGG19 architecture pretrained on the ImageNet dataset. For foot morphology classification, a fully connected network with a two-class output (high vs. low risk) was appended to the VGG19 block (256,64,32,2 neurons), with a final SoftMax output. Training was performed with leave-one-out approach and repeated 5 times for each patient. Up to 300 epochs with Early Stopping policy and a batch size of 8 were used. Code was written in Python 3 with TensorFlow 2 framework. Results are average of five independent repetitions.

## DISCUSSION

This study hypothesized that artificial intelligence could help identifying individuals with potential syndromic aortic disorders from morphological features of the forefoot. Indeed, the whole morphology of the foot is impressive in affected patients, particularly when observed from the front. However, features of the forefoot are difficult to define and measure, hence they were not explored so far to compute a score. Despite we confirmed that this is possible, further work will be needed to elucidate which combination of features the neuronal network found i.e., through explanatory AI methods.

The limited sample size complicates the application of machine learning to rare diseases. In this study we overcame this via transfer learning. More precisely, a deep neuronal network was initially trained to recognize objects from the ImageNet database that includes 1,281,167 images (e.g., animals, tools), not related to cardiovascular disorders. Then, this network was used as feature extractor, enabling the training of a smaller network specific to our dataset. Moreover, the leave-one-out method was employed for performance evaluation. However, larger datasets and studies remain essential for evaluation at population level.

Altogether, this study highlights the importance of deep learning and computer vision techniques to assist the identification of pathological phenotypes, contributing to the advancement of digital screening for rare genetic disorders, potentially enabling a broader range of healthcare professionals to perform evaluations and reducing delays for a proper management.

## Data Availability

Data are available from the corresponding authors upon reasonable request

